# Metabolomics and a Breath Sensor Identify Acetone as a Biomarker for Heart Failure

**DOI:** 10.1101/2021.05.24.21257753

**Authors:** Patrick A. Gladding, Renee Young, Maxine Cooper, Suzanne Loader, Kevin Smith, Erica Zarate, Saras Green, Silas G. Villas-Boas, Phillip Shepherd, Purvi Kakadiya, Eric Thorstensen, Christine Keven, Margaret Coe, Mia Jüllig, Edmond Zhang, Todd T. Schlegel

## Abstract

**Background:** Multiomics delivers more biological insight than targeted investigations. We applied multiomics to patients with heart failure with reduced ejection fraction (HFrEF).

**Methods:** 46 patients with HFrEF and 20 controls underwent metabolomic profiling, including liquid/gas chromatography mass spectrometry (LCMS/GCMS) and solid-phase microextraction (SPME) volatilomics in plasma and urine. HFrEF was defined using left ventricular global longitudinal strain, ejection fraction and NTproBNP. A consumer breath acetone (BrACE) sensor validated results in n=73.

**Results:** 28 metabolites were identified by GCMS, 35 by LCMS and 4 volatiles by SPME in plasma and urine. Alanine, aspartate and glutamate, citric acid cycle, arginine biosynthesis, glyoxylate and dicarboxylate metabolism were altered in HFrEF. Plasma acetone correlated with NT-proBNP (r = 0.59, 95% CI 0.4 to 0.7), 2-oxovaleric and cis-aconitic acid, involved with ketone metabolism and mitochondrial energetics. BrACE> 1.5 ppm discriminated HF from other cardiac pathology (AUC 0.8, 95% CI 0.61 to 0.92, P < 0.0001).

**Conclusion:** Breath acetone discriminated HFrEF from other cardiac pathology using a consumer sensor, but was not cardiac specific.

## Introduction

Heart failure is a common heterogeneous condition which carries significant morbidity and mortality. Despite numerous advances in heart failure management, new diagnostic tools and methods of patient stratification are needed to meet the growing demands that heart failure poses on health care providers. New tools are required to diagnose heart failure and stratify patients for treatment strategies. Metabolomics is a well-established analytical method, which involves the broad identification and quantitation of hundreds to thousands of metabolites in a single analysis. Metabolomics applied to heart failure has demonstrated its diagnostic superiority to brain natriuretic peptide (BNP) in patients with both reduced and preserved ejection fraction (1). Metabolomics has been used to reveal novel molecular pathways, altered by disease states such as heart failure (2, 3), and provides a means to monitor the effectiveness of therapies through the practice of systems pharmacology (4, 5). Despite this, however, due to numerous barriers such as standardisation, validation and platform cost, metabolomics has not been translated into clinical care (6).

Metabolomics can be applied to multiple biological tissues including plasma, urine and even breath, the latter known as volatilomics. Metabolomic profiling of volatile organic compounds (VOCs) in breath is beginning to show promise as a diagnostic tool in heart failure (7-9). For instance acetone, a product of ketone metabolism, has been demonstrated to be elevated in not only the plasma and urine of patients with heart failure but also in breath (10-13). Patterns of additional VOCs, including acetone, have been used to accurately discriminate the presence of heart failure in a non-invasive manner (14, 15). The integration of multiple sources of data or multi-omics, such as metabolomics and volatilomics applied to varying biofluids, together with genomics and protein biomarkers can provide even greater insight than any single source of data (16).

We investigated the utility of metabolomics, multi-omics and deep phenotyping in the interrogation of biological processes occurring in patients with heart failure with reduced ejection fraction (HFrEF). We then translated those findings to investigate the potential utility of a low-cost breath acetone (BrACE) sensor for predicting the presence of HF in both an inpatient and outpatient setting. In addition we evaluated these methods in identifying novel biomarkers for heart failure and ventricular arrhythmia.

## Methods

### Patients

The NanoHF study (A Novel Nanosensor array for Heart Failure diagnosis) was approved by the Northern B Health and Disability Ethics Committee (16/NTB/115) (#16/680) and Waitematā District Health Board’s IRB (#RM13458). Patients were identified from an echocardiography database, > 18 years of age, able to provide written informed consent and had previously documented signs and symptoms of heart failure with an ejection fraction between 20 and 45% on echocardiography. Exclusion criteria included diabetes mellitus (Type 1, Type 2 on insulin and/or last available HbA1c ≥ 65mmol), chronic renal impairment (eGFR <50mls/min), chronic lung disease (e.g. COPD and Asthma), and/or hospital admission within 3 months of enrolment related to exacerbation of heart failure. Heart failure was defined as a clinical syndrome accompanied by biochemical (NTproBNP; normal < 35 pmol/L and HFrEF at any age > 212 pmol/L) or mechanical (LV ejection fraction < 50%, global longitudinal strain (GLS) < 18%) abnormalities. Enrolment was enriched for patients with devices (ICD, CRT). Controls were self-reported volunteers who also underwent ECG and echocardiography.

### Hypotheses

The primary objective was to evaluate the ability of a novel breath sensor, optimised to detect acetone, and volatilomics to discriminate VOC patterns of heart failure. The secondary objective was to explore the biology and validate a metabolomic panel for heart failure.

### Biomarkers

Blood was collected using EDTA tubes. After centrifugation at 3,000 g for 5-mins, plasma was stored at −80°C before being shipped on dry ice to core lab facilities for testing. NT-proBNP was measured using a Siemens Dimension Vista assay.

### GCMS

Plasma and urine samples underwent thawing, extraction and methyl chloroformate derivatisation, as described previously (17). Gas Chromatography-Mass Spectrometry (GCMS) was used for identification and semi-quantitation of amino acids (except arginine), organic acids, and fatty acids. GCMS instrument parameters were based on Smart et al (17), using an Agilent 7890A gas chromatograph coupled to an 5975C inert mass spectrometer. Data analysis was semi-automated by using Automated Mass Spectral Deconvolution and identification software (AMDIS) against an in-house library of 165 methyl chloroformate derivatised compounds. Compounds that are not included in this library were tentatively identified using the National Institute of Standards and Technology (NIST) library. Metabolomic data were expressed as relative abundance in reference to an internal standard (DL-alanine-2,3,3,3-d_4_).

### LCMS

A targeted metabolomics approach was used to analyse plasma samples from HFrEF patients and controls. The sample preparation and analysis procedures were performed according to the AbsoluteIDQ p400 kit (Biocrates Life Sciences AG, Innsbruck, Austria) using a Thermo Q-Exactive Orbitrap LC-MS. This kit allows the measurement of 400 metabolites by UPLC-MS/MS (ultra-high performance liquid chromatography-tandem mass spectrometry) and FIA-MS (MS-based flow injection analysis). The data analysis and calculation of the metabolite concentrations analyzed by FIA were automated using MetIDQ software (Biocrates Life Sciences AG, Innsbruck, Austria).

### Volatilomics

200µl plasma and urine was used for solid-phase microextraction (SPME) volatilomics with a divinylbenzene/carboxen/polydimethylsiloxane fibre assembly. Fused silica Rxi-5Sil MS Columns were used in an Agilent 5975C Series GC/MSD using methods similar to those previously described to identify 73 VOCs (18). The original objective had been to evaluate the diagnostic utility of a novel single walled carbon nanotube sensor (SWCN) array developed at JPL/NASA Ames (19, 20). However by the time of enrolment the sensor was not ready for clinical use. We therefore tested a selection of commercially available acetone sensors including anAT6000 (NM hot wire sensor) (Greenwon, China), Keyto (Keyto, USA), Tiger LT (Photoionisation sensor) (Ion Science, United Kingdom) and Ketoscan mini (Sentech, South Korea) in a sample of cardiac inpatients and consecutive outpatients.

### Statistics

Metaboanalyst was used for pathway and multivariate analysis which was adjusted for multiplicity to reduce the false discovery rate (FDR) (21). Univariate analysis was performed using the student t-test for continuous parametric variables, a Mann Whitney U test for nonparametric and Chi square test for categorical variables. Receiver-operating characteristic curve (ROC) analysis was used to assess performance of diagnostic biomarkers by c-statistic. All tests were two-tailed and P<0.05 deemed statistically significant, except where tests for multiplicity were applied. Medcalc software version 16.8.4 was used to analyse the data. Machine learning to a validated 4 metabolite panel (1), using logistic regression (LR), random forest, decision tree and support vector machine with a 67:33 training/validation random split with tenfold cross-validation. An interactive network was generated to compare the metadata of patients using a Javascript D3 Force layout using a Pearson correlation matrix.

### Data availability

The materials, data, code, and associated protocols are available to readers with application to the corresponding author

## Results

Three hundred sixty-two patients were screened for inclusion/exclusion criteria. Sixty-six participants (46 with documented diagnosis of heart failure and 20 self-reported healthy volunteers) were enrolled in the study, with written informed consent. Baseline characteristics are outlined in Table 1. Twenty-seven (59%) of the heart failure patients had an ischaemic cardiomyopathy and 19 (41%) had either an ICD (n=14) or CRTD (n=5). Ten (71%) of ICDs were implanted for primary prevention. Heart failure patients were older and had a higher percentage of males than controls. Seventeen (37%) of heart failure patients had normal NTproBNP, indicating biochemical HF recovery (HFrec). Mean NYHA status was II.

**Table 1.** Baseline Characteristics

|  | HF<br>n=46 | Controls<br>n=20 | P value |
| --- | --- | --- | --- |
| Age, mean (SD) | 68 (8) | 52 (9) | 5x10<sup>-9</sup> |
| Male, n (%) | 41 (89) | 10 (50) | 0.0006 |
| European | 29 (63) | 16 (80) | 0.18 |
| AF | 10 (22) | 0 (0) | N/A |
| HTN | 21 (46) | 0 (0) | N/A |
| T2DM | 9 (20) | 0 (0) | N/A |
| ACEi/ARB | 37 (80) | 0 (0) | N/A |
| Beta blocker | 39 (85) | 0 (0) | N/A |
| MRA | 14 (30) | 0 (0) | N/A |
| Statin | 29 (63) | 0 (0) | N/A |
| Furosemide | 10 (22) | 0 (0) | N/A |
| EF bp mean (SD) | 39% (10) | 57% (5) | 8x10<sup>-9</sup> |
| GLS | -13% (0.04) | -21% (0.05) | 3x10<sup>-8</sup> |
| NTproBNP (pmol/L) | 115 (124) | 8 (10) | 0.0002 |

AF = atrial fibrillation, HTN = hypertension, T2DM = type 2 diabetes, ACEi = angiotensin converting
enzyme inhibitor, ARB = angiotensin receptor blocker, MRA = mineralocorticoid receptor antagonist,
EF bp = ejection fraction by Simpson's biplane, GLS = global longitudinal strain.

### Metabolomics

28 metabolites across all diagnostic definitions of heart failure were identified by GCMS which met FDR (Table 2). Numerous of these were either directly part of or indirectly linked to the citric acid cycle and mitochondrial metabolism. By univariate analysis, isocitric acid had the highest AUC 0.84, 95% CI 0.73 to 0.92. 35 metabolites were identified by LCMS which fulfilled the FDR (Table 3). Most notably these included symmetric dimethyl arginine, creatinine, arginine and kynurenine as well as numerous phosphatidylcholines, sphingomyelins, lysophosphatidylcholines, two cholesteryl esters and one triglyceride (55:9). Nineteen common metabolites were detected by both GCMS and LCMS allowing for analytical method comparisons.

**Table 2.**
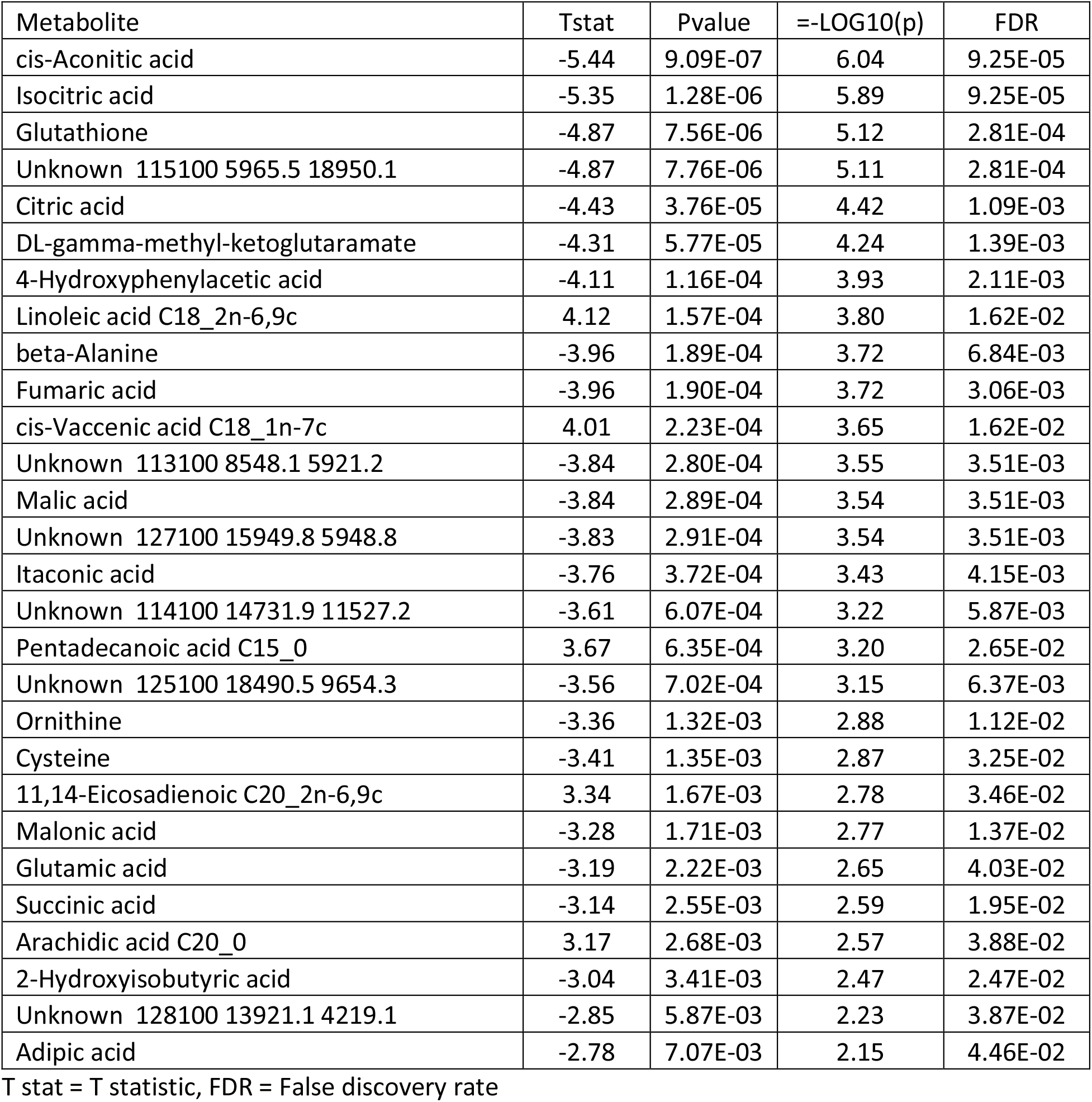
Metabolites identified by GCMS as significantly different in heart failure

| Metabolite | Tstat | Pvalue | =-LOG10(p) | FDR |
| --- | --- | --- | --- | --- |
| cis-Aconitic acid | -5.44 | 9.09E-07 | 6.04 | 9.25E-05 |
| Isocitric acid | -5.35 | 1.28E-06 | 5.89 | 9.25E-05 |
| Glutathione | -4.87 | 7.56E-06 | 5.12 | 2.81E-04 |
| Unknown 115100 5965.5 18950.1 | -4.87 | 7.76E-06 | 5.11 | 2.81E-04 |
| Citric acid | -4.43 | 3.76E-05 | 4.42 | 1.09E-03 |
| DL-gamma-methyl-ketoglutaramate | -4.31 | 5.77E-05 | 4.24 | 1.39E-03 |
| 4-Hydroxyphenylacetic acid | -4.11 | 1.16E-04 | 3.93 | 2.11E-03 |
| Linoleic acid C18_2n-6,9c | 4.12 | 1.57E-04 | 3.80 | 1.62E-02 |
| beta-Alanine | -3.96 | 1.89E-04 | 3.72 | 6.84E-03 |
| Fumaric acid | -3.96 | 1.90E-04 | 3.72 | 3.06E-03 |
| cis-Vaccenic acid C18_1n-7c | 4.01 | 2.23E-04 | 3.65 | 1.62E-02 |
| Unknown 113100 8548.1 5921.2 | -3.84 | 2.80E-04 | 3.55 | 3.51E-03 |
| Malic acid | -3.84 | 2.89E-04 | 3.54 | 3.51E-03 |
| Unknown 127100 15949.8 5948.8 | -3.83 | 2.91E-04 | 3.54 | 3.51E-03 |
| Itaconic acid | -3.76 | 3.72E-04 | 3.43 | 4.15E-03 |
| Unknown 114100 14731.9 11527.2 | -3.61 | 6.07E-04 | 3.22 | 5.87E-03 |
| Pentadecanoic acid C15_0 | 3.67 | 6.35E-04 | 3.20 | 2.65E-02 |
| Unknown 125100 18490.5 9654.3 | -3.56 | 7.02E-04 | 3.15 | 6.37E-03 |
| Ornithine | -3.36 | 1.32E-03 | 2.88 | 1.12E-02 |
| Cysteine | -3.41 | 1.35E-03 | 2.87 | 3.25E-02 |
| 11,14-Eicosadienoic C20_2n-6,9c | 3.34 | 1.67E-03 | 2.78 | 3.46E-02 |
| Malonic acid | -3.28 | 1.71E-03 | 2.77 | 1.37E-02 |
| Glutamic acid | -3.19 | 2.22E-03 | 2.65 | 4.03E-02 |
| Succinic acid | -3.14 | 2.55E-03 | 2.59 | 1.95E-02 |
| Arachidic acid C20_0 | 3.17 | 2.68E-03 | 2.57 | 3.88E-02 |
| 2-Hydroxyisobutyric acid | -3.04 | 3.41E-03 | 2.47 | 2.47E-02 |
| Unknown 128100 13921.1 4219.1 | -2.85 | 5.87E-03 | 2.23 | 3.87E-02 |
| Adipic acid | -2.78 | 7.07E-03 | 2.15 | 4.46E-02 |

T stat = T statistic, FDR = False discovery rate

**Table 3.**
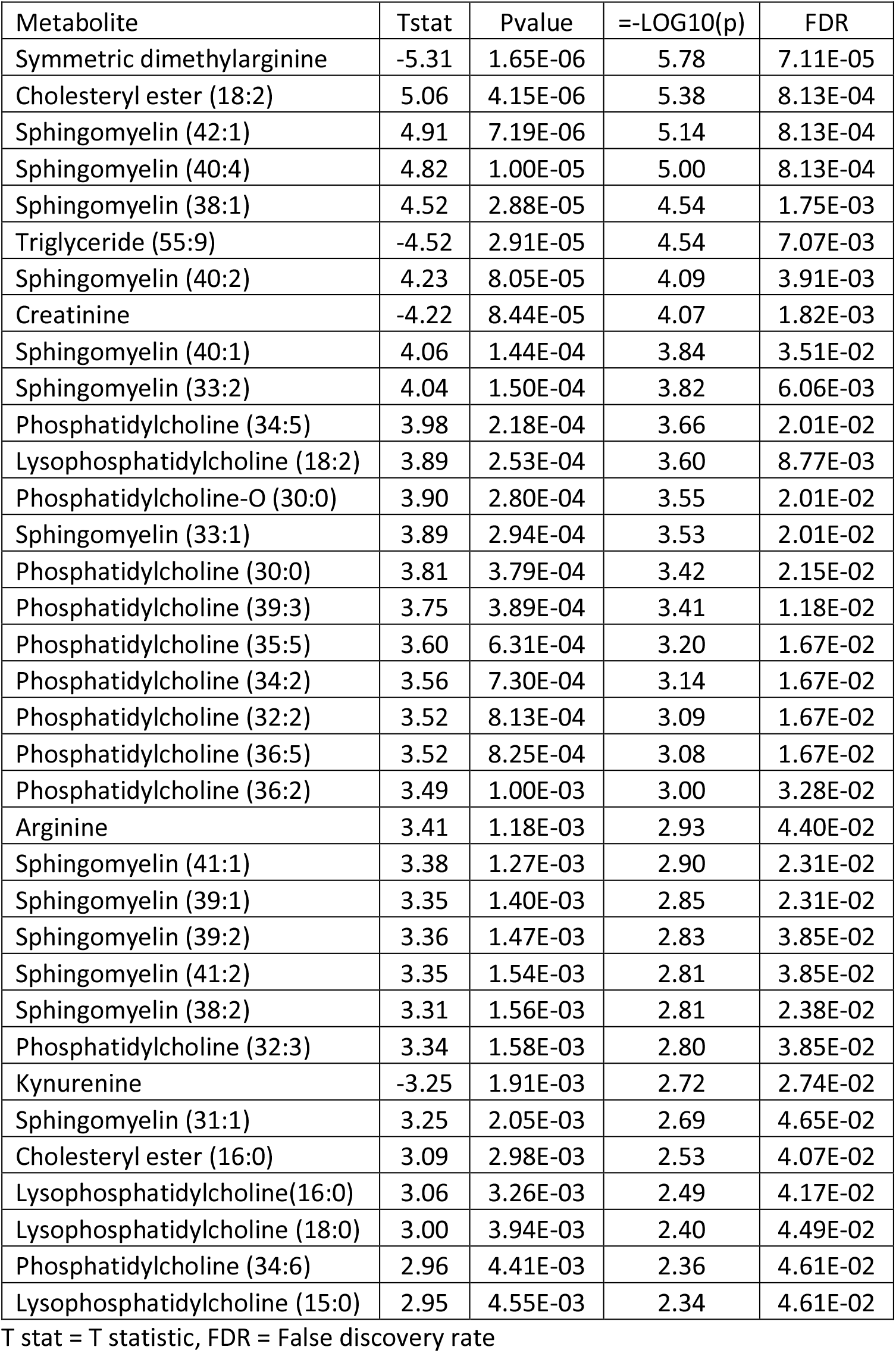
Metabolites identified by LCMS as significantly different in heart failure

Using a previously validated panel of metabolites (histidine, phenylalanine, spermidine, and phosphatidylcholine C34:4) (1) and LR, an AUC of 0.92 was achieved, accuracy 0.85, precision 0.79, recall 0.85, F1 score 0.82, compared to NTproBNP AUC 0.93, 95% CI 0.83 to 0.98, for the discrimination of heart failure.

### Volatilomics

Only one volatile, acetone, reached significance by the stringent FDR used (Table 4, Figures 1 and 2), however several common VOCs were identified in both plasma and urine (t-test, P<0.05) which have previously been associated with heart failure. These included pentane, 2-butanone, and 2-pentanone.

**Table 4.** VOC identified by SPME as significantly different in heart failure

|  | Tstat | Pvalue | =-LOG10(p) | FDR |
| --- | --- | --- | --- | --- |
| Acetone | -6.0327 | 9.80E-08 | 7.0088 | 1.08E-05 |

T stat = T statistic, FDR = False discovery rate

**Figure 1.**
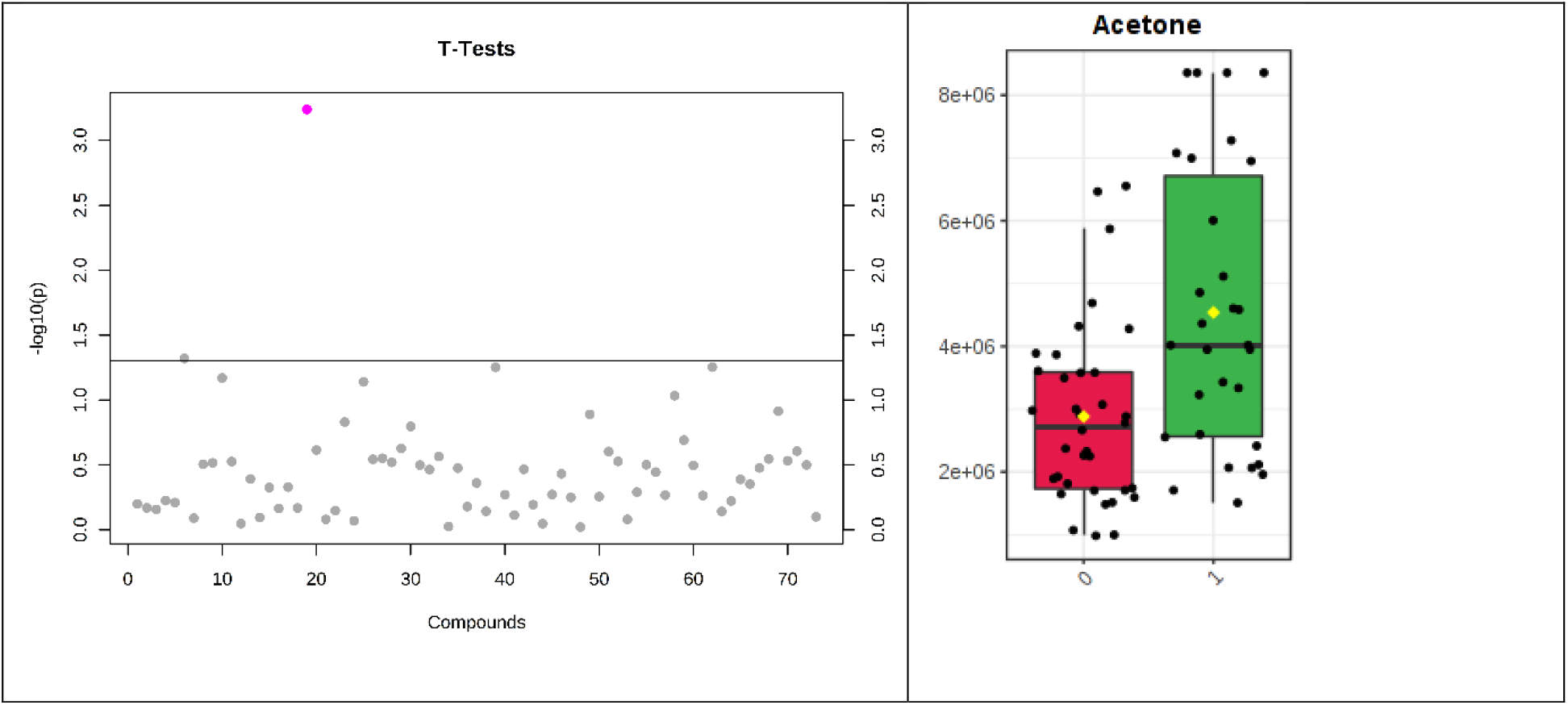
SPME plasma NTproBNP ≥ 35pmol/L Log10 transformed t test p values for individual metabolites with p value threshold for multiplicity (horizontal line) (left). Single VOC reaching statistical significance (right).

**Figure 2.**
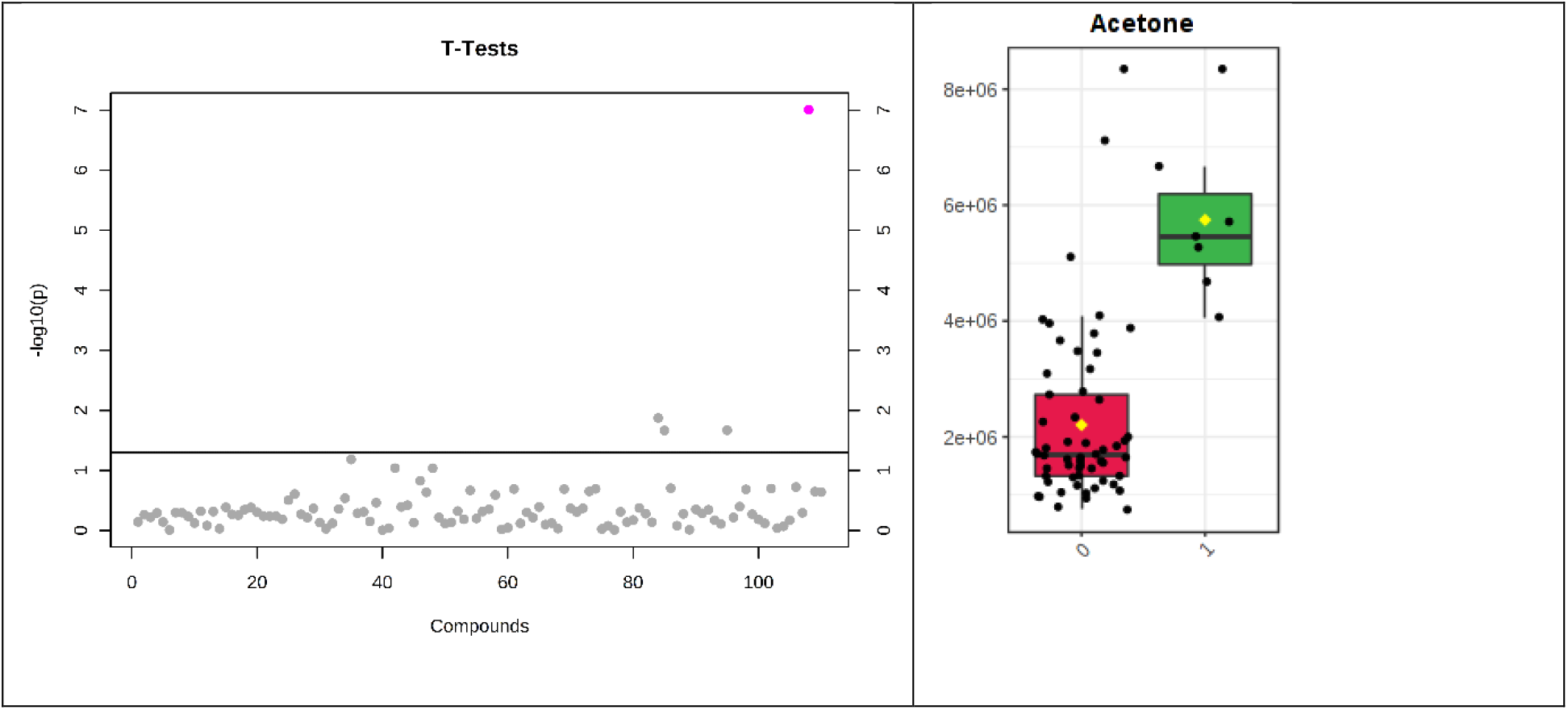
SPME urine NTproBNP > 211pmol/L Log10 transformed t test p values for individual metabolites with p value threshold for multiplicity (horizontal line) (left). Single VOC reaching statistical significance (right).

Of the three consumer sensors tested only one had the sensitivity, reproducibility and accuracy to identify breath acetone (BrACE) differences in HF patients versus controls in the clinical validation (n=61, inpatient/outpatients n=32/29). BrACE had an AUC of 0.8, 95% CI 0.61 to 0.92 for discriminating heart failure in 12 (41%) cardiac outpatients with compensated heart failure. BrACE was higher in acute decompensated heart failure (HFpEF and HFrEF) versus compensated patients (median 2.6 versus 1.6 ppm, P = 0.046). However in a sample of breathless inpatients (n=12) with noncardiac pathology, BrACE was 3.2 +/-2.7 ppm (mean/SD) (pneumothorax, pneumonia, lung malignancy, uncontrolled diabetes), reflecting increases in ketone body production due to physiological stress (Central Illustration).

### Pathway and network analysis

Metabolites from the citric acid cycle dominated the pathway analysis, with other pathways identified including arginine, alanine, aspartate and glutamate biosynthesis, as well as glyoxylate and dicarboxylate metabolism. Networks showed multiple hubs, including GLS with a high degree of betweenness centrality https://projects.interacta.io/theranostics/. Plasma acetone correlated with NT-proBNP (r = 0.59, 95% CI 0.4 to 0.7), triacylglycerol (55:9), 2-oxovaleric and cis-aconitic acid, involved with ketone metabolism and mitochondrial energetics.

## Discussion

In this study we took a multiomics approach using urinary and plasma metabolomics, volatilomics, biomarkers and DNA sequencing to deeply phenotype patients with HFrEF. Our main objectives were to evaluate the volatilome of patients with heart failure, using plasma and urine as a surrogate for breath, and then validate our results using a widely available breath acetone sensor. We also used untargeted and targeted metabolomics to validate a 4 metabolite panel previously shown to be diagnostic for HFrEF (1).

Acetone was first identified as a potential biomarker in the breath of heart failure patients in 1995 by Kupari et al (11). Breath acetone (BrACE) has then subsequently been shown to have a similar diagnostic accuracy to BNP, with acetone concentrations reflecting heart failure severity (22). This was reinforced by a longitudinal study showing elevated BrACE concentration correlated with PCWP (BrACE >1.05 ppm was associated with PCWP ≥ 18 mmHg, AUC 0.72) (23) and was associated with a poorer prognosis (24). Similarly urine acetone has been shown to correlate with heart failure (25) and its severity, determined by NYHA class and echocardiography (12).

In our study our intention had been to validate BrACE using a SWN sensor array, capable of discriminating VOC patterns similar to mass spectrometry; however, the sensor was not ready for clinical use. A previous study by Samara et al, at the Cleveland Clinic, used SIFT-MS (Syft technology, New Zealand) and discrimimant analysis, a form of machine learning, to identify a breath fingerprint for heart failure (8). This included acetone, pentane and other ion peaks which have subsequently been validated along with isoprene (26), 2-pentanone and 1-butanol in a detailed study by Biagini et al (15). Biagnini similarly proposed the use of BrACE as a biomarker and potential monitor for heart failure, especially as it reduced three fold in response to in-hospital treatment for acute decompensation. We similarly showed that not only is BrACE a potential diagnostic for heart failure, but is also at lower concentration in the stable compensated setting. We have for the first time demonstrated the measurement of BrACE with a low cost consumer sensor (Ketoscan mini, Senentech, Korea), with sufficient sensitivity, precision and accuracy to measure acetone < 2ppm (+/-1ppm < 5ppm), in the healthy range (27). The main limitations are that BrACE levels are confounded by prolonged starvation, uncontrolled diabetes, ketogenic diets and the use of lozenges/gum containing menthol. Despite the SWN sensor being unavailable for clinical use we are continuing to work on a sensor array, using consumer-available sensors (28).

Metabolomics has provided a wealth of information about not only the mechanisms underlying heart failure but also suggested therapies. Ketone bodies, such as acetoacetate and beta-hydroxybutyrate (βOH), are one of the many energy substrates for the heart, inclusive of glucose, branched chain amino acids and free fatty acids. It is believed that ketone body formation from the liver is an early adaptive response to heart failure. Though debate still exists around whether this is a positive or negative adaptive mechanism a recent knockout model would suggest the former (29). Deletion of succinyl-CoA:3-ketoacid-CoA transferase 1 (SCOT) In a mouse model increased circulating ketones and reduced the cardiac inflammasome preventing heart failure caused by increased afterload (30). Metabolomics has also shown that beta-hydroxybutyrate, acetone and succinate correlate with myocardial energy expenditure measured by echocardiography (kcal/min) and act independently to treatment with angiotensin converting enzyme inhibitor, β-receptor blockers, diuretics and statins (31). This would suggest a mechanism independent of these traditional therapies, which is open to modification. In fact empagliflozin, an SGLT2 inhibitor, increases ketone body formation, but not cardiac substrate utilisation, which may in part explain its mechanism of action (32, 33). Similarly nutritional interventions such as βOH, which can be taken orally, may have a role in heart failure. βOH, given as an infusion, has been shown to increase overall energy production without compromising glucose or fatty acid metabolism, albeit without increasing cardiac efficiency (34, 35).

In our study untargeted GCMS/LCMS metabolomics revealed a number of pathways altered in heart failure, with mitochondrial metabolism at the forefront. Although standardisation has been an issue with metabolomics, we have shown a 4-panel metabolite profile, using an LCMS kit, produces a reproducible result, equivalent to the diagnostic capacity of NTproBNP (1). LCMS identified arginine and symmetric dimethylarginine in the kynurenine pathway, previously implicated in heart failure (1, 36). Tryptophan and the kynurenine pathway are intimately linked to NAD+ production and supplementation with NAD donors, such as nicotinamide riboside, have been suggested as a potential therapeutic in heart failure (37). As ketone utilisation and mitochondrial function are intimately linked, the measurement of BrACE opens up the possibility of a real time probe of body metabolism, as either a screening tool for diagnosis or monitoring of pharmacological therapy or nutritional interventions with mitochondrial therapies. Mitochondrial therapies might be particularly effective in titin truncation cardiomyopathy (38) more so than in other cardiomyopathies (39), though this is yet to be fully elucidated.

## Limitations

This study was limited by its small sample size. Moreover, results that were not prespecified can only be considered exploratory. The GCMS analysis used an in-house non-quantitative method, which may not be reproducible elsewhere. However the p400 LCMS kit did provide quantitation on a commonly available, well validated mass spectrometry platform which has also previously been used in heart failure (1). Although the sample preparation steps for LCMS may limit its application in a clinical laboratory, other methods such as NMR may provide the scalability and utility for clinical use. 700 MHz NMR results are awaited for this study, which will provide further quantitation of metabolites identified by both GCMS and LCMS.

## Data Availability

Data is available on request

## Acknowledgements

Health Research Council of New Zealand Explorer Grant 16/680, Auckland Regional Tissue Bank. Nikita Rokotyan, data visualisation. Umit Holland and Victoria Anderson for study co-ordination.

## Central Illustration

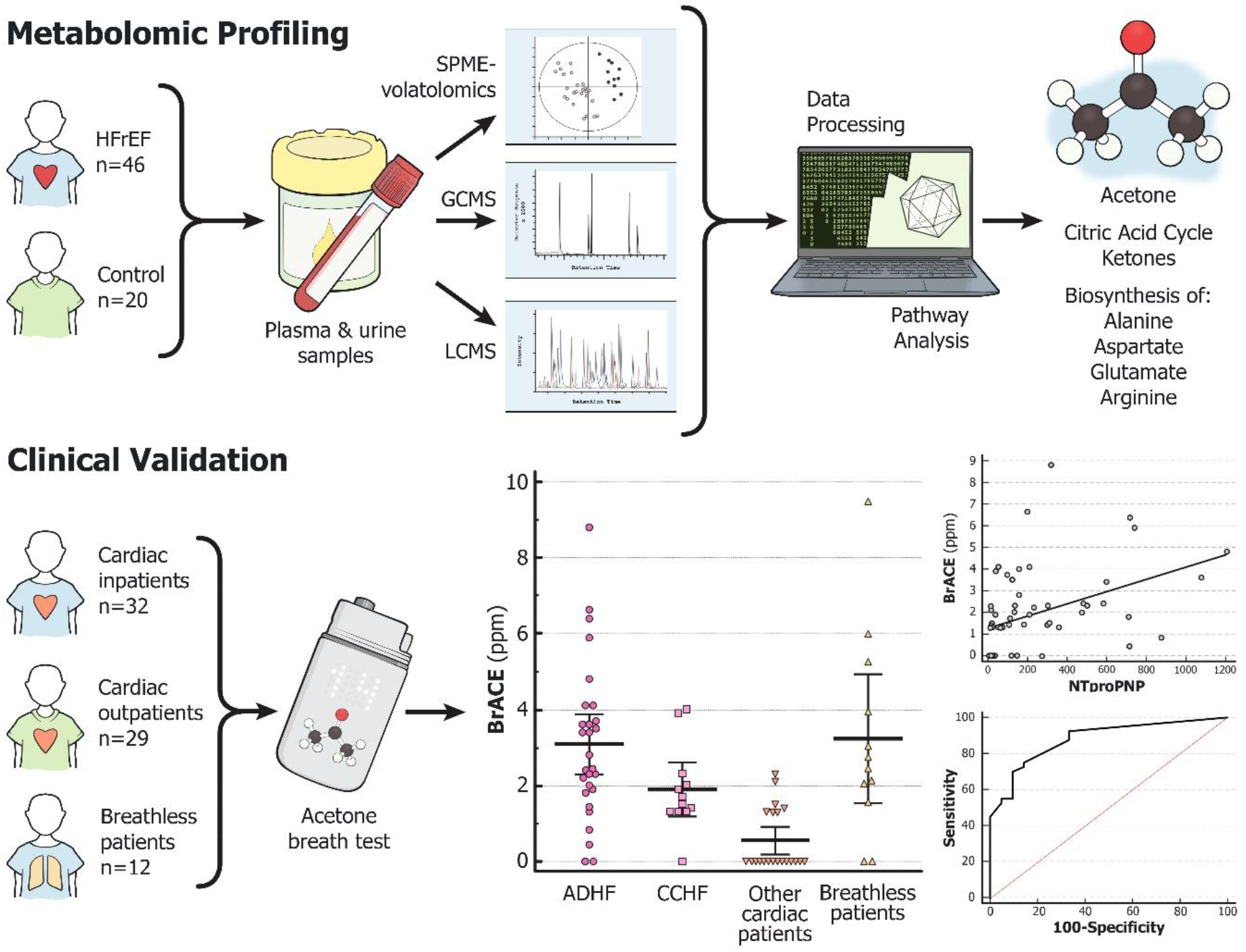

ADHF = acute decompensated heart failure, CCHF = chronic compensated heart failure, SPME = Solid-phase microextraction, GCMS = Gas chromatography mass spectrometry, LCMS = Liquid chromatography mass spectrometry

## Notes

### Competing Interest Statement

The authors have declared no competing interest.

### Author Declarations

Northern B Health and Disability Ethics Committee 16/NTB/115, 16/680 and Waitematā District Health Board IRB RM13458

